# Validation of a MALDI-TOF MS method for SARS-CoV-2 detection on the Bruker Biotyper and nasopharyngeal swabs. A Brazil - UK collaborative study

**DOI:** 10.1101/2023.01.09.23284367

**Authors:** Otávio A. Lovison, Raminta Grigaitė, Fabiana C. Z. Volpato, Jason K. Iles, Jon Lacey, Fabiano Barreto, Sai R. Pandiri, Lisiane da Luz R. Balzan, Vlademir V. Cantarelli, Afonso Luis Barth, Andreza F. Martins, Ray K. Iles

## Abstract

We had developed a MALDI-TOF mass spectrometry method for detection of SARS-CoV-2 virus in saliva-gargle samples using Shimadzu MALDI-TOF mass spectrometers in the UK. This was validated in the USA to CLIA-LDT standards for asymptomatic infection detection remotely via sharing protocols, shipping key reagents, video conference and data exchange. In Brazil, more so than in the UK and USA, there is a need to develop non-PCR dependent rapid affordable SARS-CoV-2 infection screening tests, which also identify variant SARS-CoV-2 and other virus infections. Travel restrictions necessitated remote collaboration with validation on the available Clinical MALDI-TOF – the Bruker Biotyper (microflex® LT/SH) – and on nasopharyngeal swab samples, as salivary gargle samples were not available. The Bruker Biotyper was shown to be almost log10^3 more sensitive at detection of high molecular weight spike proteins. A protocol for saline swab soaks out was developed and duplicate swab samples collected in Brazil were analysed by MALDI-TOF MS. The swab collected sample spectra varied from that of gargle-saliva in three additional mass peaks in the mass region expected for IgG heavy chains and human serum albumin. A subset of clinical samples with additional high mass, probably Spike-related proteins, were also found. Spectral data comparisons and analysis, subjected to machine learning algorithms in order to resolve RT-qPCR positive from RT-qPCR negative swab samples, showed a 78% agreement with RT-qPCR scoring for SARS-CoV-2 infection.

## 1. Introduction

Brazil is an upper-middle-income country with 210 million inhabitants in a large territorial area. There is substantial socioeconomic heterogeneity among its five macro-regions which is reflected in the health services, including the availability of hospital beds and trained healthcare workers [1]. In 2021, the Ministry of Health of Brazil published the ‘National Plan to Expand Testing for COVID-19 [2]. To address the objectives of the plan, clinical and research laboratories have expanded their routine to perform RT-qPCR and high-throughput automated testing that was implemented mainly in reference centres. Despite the effort made, the laboratories became overloaded, while having to deal with SARS-CoV-2 testing kits and laboratory consumables shortages [3].

In addition, insufficient sanitary regulation, inadequate orientation for the population, official communication not based on scientific evidence, people refusing to wear masks, not observing social distancing, and believing on not proven ‘miracle pills (widespread self or prescribed medication with chloroquine/hydroxychloroquine and ivermectin)’, the pandemic followed an uncontrolled rhythm reaching 82.869 notified cases and 4.211 deaths in a single day [4]. The intra-hospital mortality was high and many people died in the absence of ICU beds and respiratory support [1]. By the end of September 2022, Brazil had 34.672.524 cases and 686.036 deaths by COVID-19 [4]. In this scenario with limited resources, the development of rapid and cost-effective screening tests is needed to manage this or new viral pandemics.

The use of MALDI-TOF mass spectrometry in clinical bacteriology has made a substantial impact on the cost of microbiological testing widening provision of them [5], [6]. Indeed, the use of this technology could simultaneously cut costs and increase SARS-CoV-2 screening, as well as other viruses, which is extremely important to improve healthcare in Brazil.

A MALDI-TOF mass spectrometry technique for SARS-CoV-2 detection had been developed by Prof. Ray Iles and his team in the UK [7, 8]. In this context, an international collaboration was formed between MAP Sciences UK and institutions in Southern Brazil, to trial and validate the methodology using Bruker Biotyper (microflex® LT/SH) and nasopharyngeal swab samples. In order to achieve the aim, the UK team at MAP Sciences established and validated the performance characteristics of Brazilian equipment against the Shimadzu 8020 MALDI – TOF mass spectrometer; and the Brazilian team prepared collected swab samples and performed analysis for which RT-qPCR results had been obtained. Data analysis was performed by MAP Sciences.

## 2. Materials and Methods

### 2.1. Samples

Pseudotype lentiviral constructs expressing the SARS-CoV-2 were obtained from Professor Nigel Templeton of the Pseudotype Unit at the University of Essex, UK. In addition, acetone extracted culture media from Hek293 cells infected with SARS-CoV-2 (original UK isolation of Alpha variant) was obtained from Jonathan Heeney of the Laboratory of Viral Zoonotics, Department of Veterinary Medicine, at the University of Cambridge, UK.

Gargle samples from 222 individuals from Bedford, UK were obtained from volunteer asymptomatic individuals in Bedford UK and collected by MAP Sciences researchers between 7th of July 2020 and the 14th of February 2022. In addition, 37 gargle-saliva samples for COVID-19 patients, who had recovered and were 3 months after discharge from the COVID respiratory care wards of Papworth Hospital, were collected by Dr Helen Baxendale and colleagues of Cambridge Universities Hospital NHS trust, Cambridge UK.

Nasopharyngeal swab samples were collected in May, 2021 from Porto Alegre, South Brazil, to the MALDI-TOF validation protocol and 248 were randomly selected for this study. These samples were previously submitted to RT-qPCR according to CDC protocol. Among them, 100 were considered positive (Ct<40) and 148 negative (Ct>40) for SARS-CoV-2 by RT-qPCR. In total 92.3% (229/248) of the patients reported respiratory symptoms.

### 2.2. MALDI-TOF Mass Spectrometry

In the UK a Shimadzu 8020 MALDI TOF owned by MAP Sciences and a loaned Bruker Biotyper, (microflex® LT/SH, Bruker, Coventry, UK) from Bruker, UK, were available for the study. The Brazilian research laboratory at Porto Alegre runs a Bruker Biotyper (microflex® LT/SH) which was used to profile isolated virion envelope proteins.

The initial development for viral detection by MALDI-TOF mass spectrometry was designed for gargle samples and used a Shimadzu 8020 MALDI-TOF mass spectrometer. Thus, the first experiments were a direct comparison of a Bruker Biotyper on the same pseudotype SARS-CoV-2, extracted cultured SARS-CoV-2 and the same gargle-saliva samples analysed by a Shimadzu 8020 MALDI-TOF. This was conducted in the UK at MAP Sciences in the UK [7].

The second consideration was adapting the pre-existing sample processing protocols for gargle saliva samples to utilise the nasopharyngeal swab sample collected in Brazil. Upon the sample collection, swabs were inserted and stored in 1 mL of saline. Thus, we had to work with a limited volume of a sample to empirically identify the most reliable method for viral protein extraction and enrichment prior to mass spectrometry. As opposed to the gargle sample, no filtration was necessary as large particulate matter was not a major interfering factor. The following method was applied; 500 µl of the soak-out swab saline was mixed with 500 µl ice-cold (4°C) acetone. Samples were placed in a centrifuge and span at 16000 RCF, at 4°C, for 30 minutes. The resulting pellet was reconstituted in 50ul LBSD-X buffer (MAP Sciences, UK) with 20 mM tris(2-carboxyethyl)phosphine (TCEP) and it was plated in duplicates in a sandwich technique after 15 minutes with a 15 mg/mL concentration of Sinapinic acid (SA) matrix.

Initially, calibration of the Bruker Biotyper was set using the established and recommended calibrants and fitting curve for biotyping as provided and recommended by Bruker. Subsequently, in the UK only, both the Shimadzu 8020 MALDI-TOF mass spectrometer and Bruker Biotyper (microflex® LT/SH, Bruker, Coventry, UK) were calibrated using a 2-point calibration of 2 mg/mL bovine serum albumin (33,200 m/z and 66,400 m/z). Mass spectral data were generated in a positive ion, linear mode. For the Bruker Biotyper, the laser power was set at 65% and the spectra were generated at a mass range between 10,000 to 200,000 m/z; pulsed extraction was set to 1400 ns.

### 2.4. Bioinformatics

Data files were quality-checked, identifying reference peak in a region of 10,000 - 11,900 m/z as previously described [8]. Spectral data was preprocessed by smoothing (single-cycle, Gaussian smoothing method with a window size of 150 m/z, and a baseline correction). Peak picking as a maximum height was performed with a 1% deviation from the predefined range of key peaks. Peaks that were not consistent throughout the dataset were removed, leaving a total of 24 features. Missing peaks on spectra were imputed with 0.01.

Based on our previous studies on gargle-saliva samples and on antibody analysis, peaks were assigned to Immunoglobulin chains and, where present, SARS-CoV-2 proteins [9-11].

In an undirected peak/mass spectra comparison approach, machine learning (ML) was applied as follows. Prior to algorithm training, the data were checked for normality and a 60-40 % test train split was made. Decision tree-based algorithms performed best and the grid search hyperparameter tuning was applied on Random Forest and Extra Tree Classifier with fitting the best parameters. Additionally, the Out of Bag Score was used on the random forest for confidence in the accuracy of the predictions.

## 3. Results

Comparing the performance of the Bruker Biotyper with the Shimadzu 8020 MALDI-TOF on pseudotype Lentivirus expressing the SARS Spike protein complex, we found that the peak masses were consistently higher by 3-4% in mass and the intensity of the higher mass peaks were also higher in recorded intensity by log10^3 (see Figure 1).

**Figure 1.**
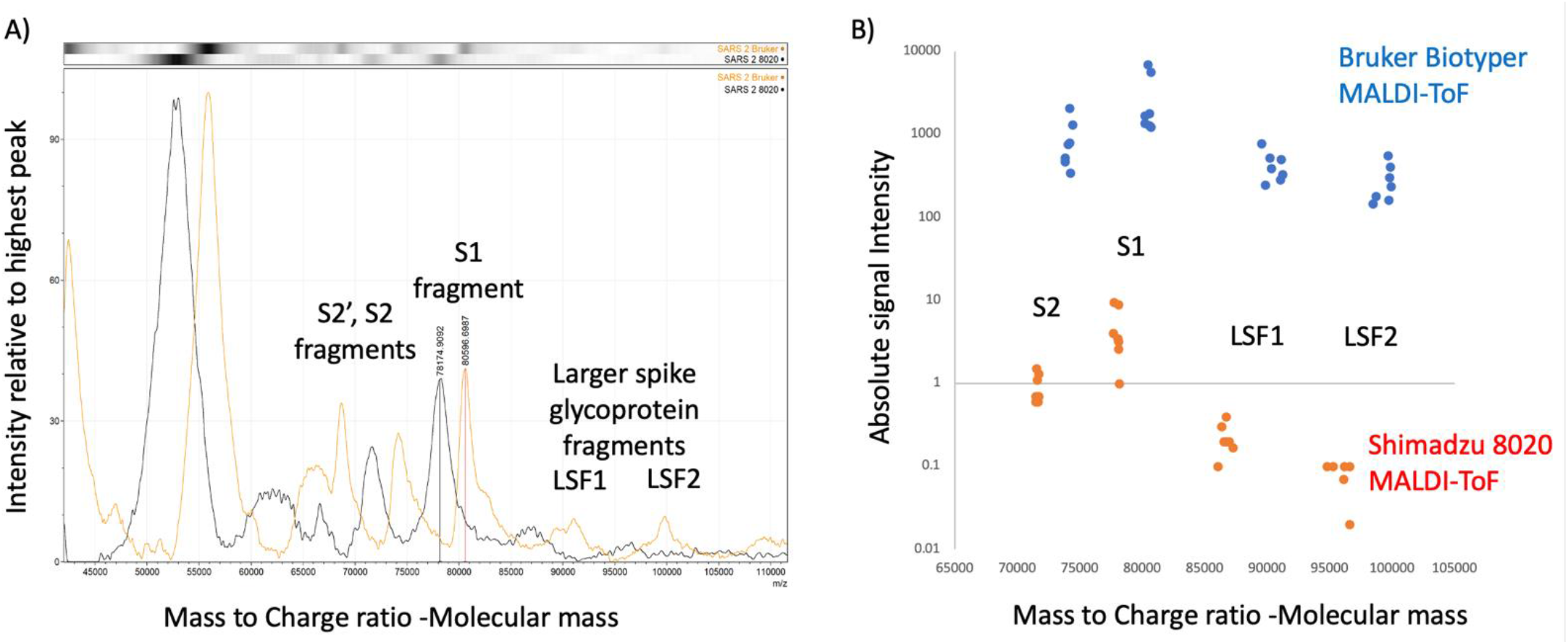
Calibration and Intensity differences Bruker Biotyper versus Shimadzu 8020 mass spectra: A) comparison of peak masses found for Lentiviral pseudotype constructs expressing the Envelope spike protein of the SARS CoV-2 Spike gene. S1 - Spike protein fragment 1; S2 - Spike protein fragment 2; B) comparison of mass and intensity difference for eight LentiViral pseudotype constructs expressing the Corona Virus’ respective Envelope Spike genes (SARS-CoV-1, SARS-CoV-2, MERS, NL3, OC43, 229E, HKU1).

By recalibrating the Bruker using bovine serum albumin (BSA) as a two-point calibrant (single and double charged BSA) the masses on subsequent spectral analysis of *in vitro* cultured SARS-CoV-2 (Alpha) matched (see figure 2).

**Figure 2.**
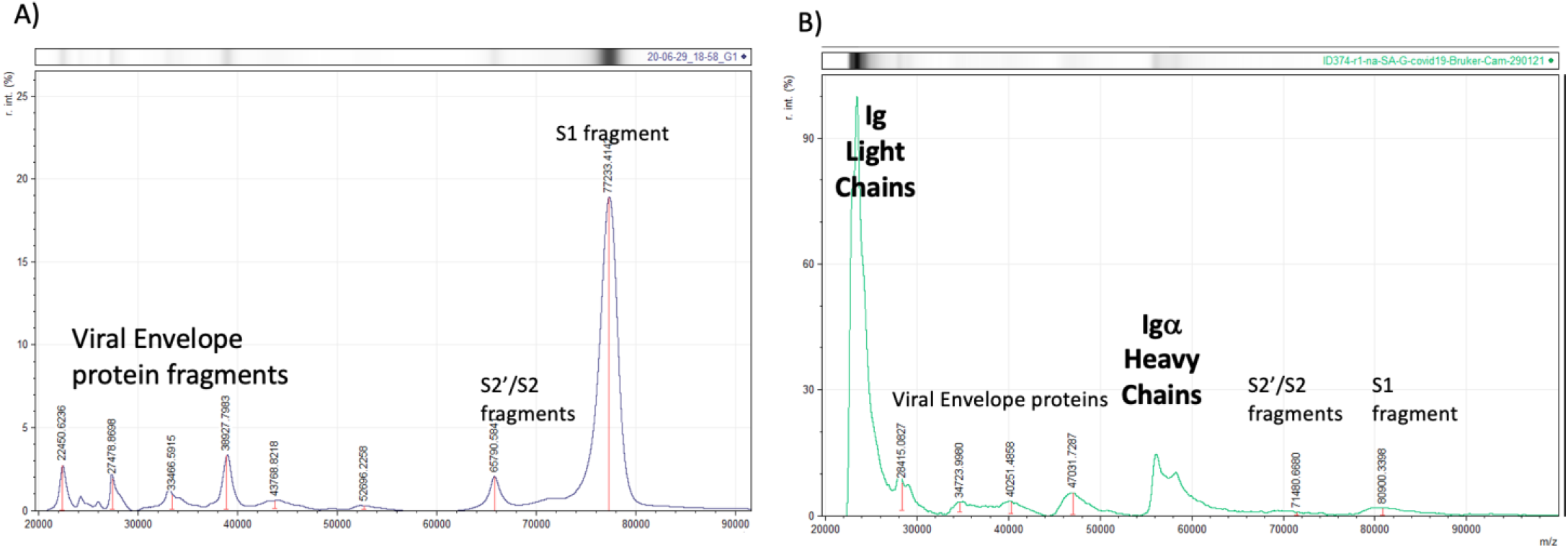
Bruker Biotyper Mass spectral profiles after higher mass drift correction by two-point (H+ and 2H+) calibrating with BSA: A) In Vitro spectra - Culture supernatant, UK isolate of COVID-19 (first wave alpha) grown on HEK293 cells. B) Clinical spectra - second wave gargle-saliva from asymptomatic RT-qPCR positive – Male 60’s.

Looking at gargle-saliva samples, where calibration had been corrected to the method employed in development of the Shimadzu 8020 SARS-CoV-2 assay; the Bruker Biotyper produced spectra with matching peaks to that generated by the Shimadzu 8020 MALDI-TOF MS. However, these were between 50 and 100 times higher in intensity on the Bruker Biotyper MALDI spectra than the same gargle-saliva samples run on the Shimadzu 8020 MALDI-TOF MS (see Figure 3). Consequently, a higher cut off value would be needed in distinguishing positive and negative samples.

**Figure 3.**
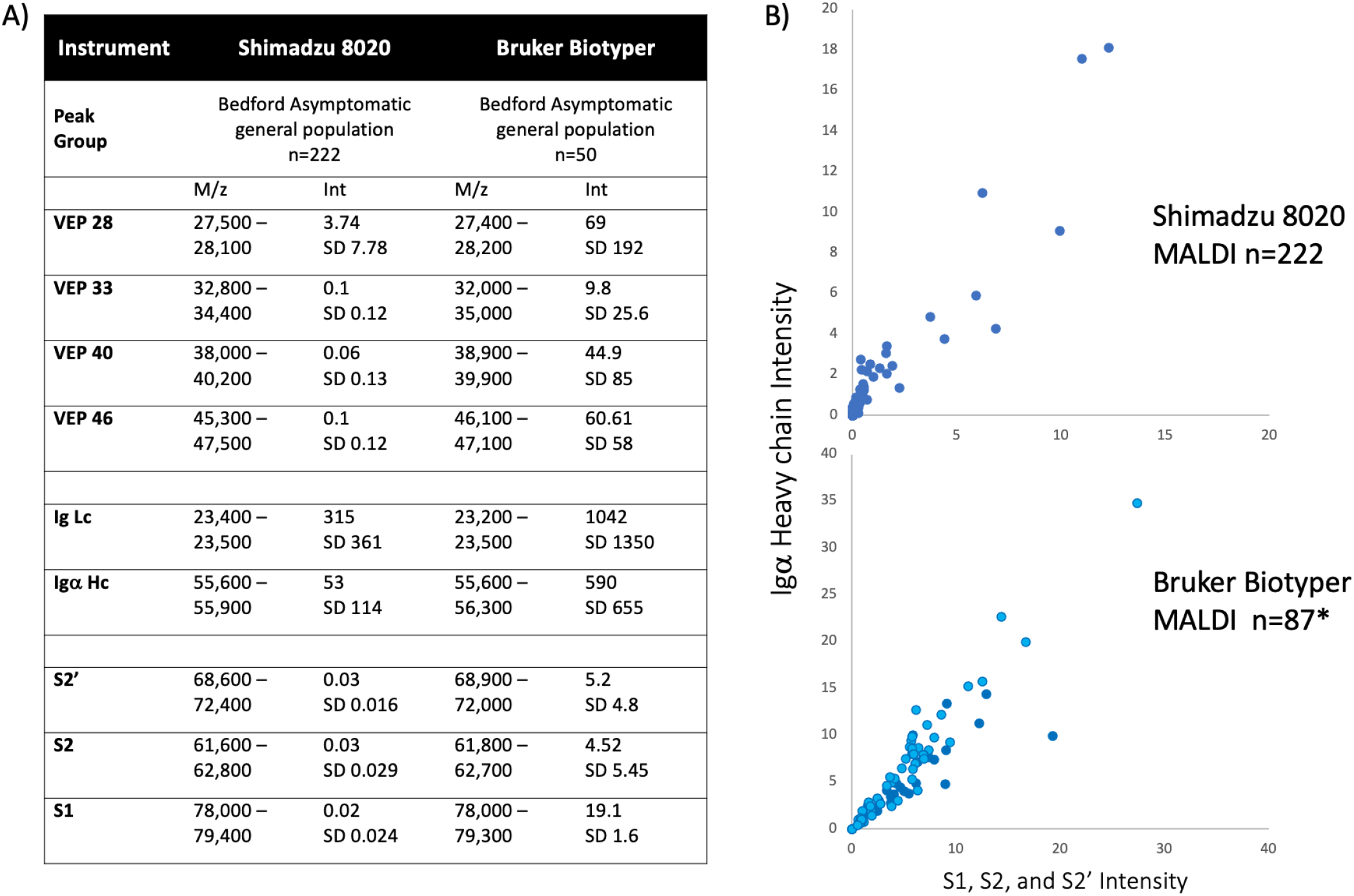
Comparison of spectral data generated from the Shimadzu 8020 and Bruker Biotyper MALDI-TOF mass spectrometer on gargle saliva samples A) tabulation of mass and intensity of viral envelope, immunoglobulin and spike fragments. VEP - Viral envelope protein; Lc - Light chain; Hc - Heavy chain; S1 - Spike protein fragment 1; S2 - Spike protein fragment 2; S2’ - Spike protein fragment 2’ B) plot of IgA heavy chain intensity versus spike protein intensity for the Shimadzu 8020 (upper panel) and Bruker Biotyper mass spectrometers. * refers to an additional 37 convalescent COVID-19 patient gargle samples added to the plot (dark blue circles).

Among 248 Brazilian swab samples, 215 produced spectra that met QC criteria (86.7%). Of these 79 were SARS-CoV-2 RT-qPCR positive and 136 were RT-qPCR negative. These spectra revealed slightly different patterns. First of note was that samples run in Brazil had a mass shift which can be attributed to machine calibration employed for bacterial biotyping not precisely matching high mass protein mass fitting. At high masses the line fitting dependent average mass shift was a 700-4000m/z higher value than those recorded in the UK values for immunoglobulin chains (Ig light chains approx. 23,000m/z) and SARS-CoV-2 viral envelope proteins (highest detected being approx. 100,000m/z). This was not corrected for *in silico* as such manipulation may have introduced a bias in ML interpretation of the results.

Significantly the presence of three new peaks corresponding to the expected masses of Human Serum Albumin (HSA) and Igγ1 and Igγ3 heavy chains, was a common finding in nasopharyngeal swab samples and very rarely seen in gargle samples. This was in addition to Ig light chains and Igα heavy chains, which were the dominant if only immunoglobulin found in saliva gargle samples (figure 4). In a subset of nasopharyngeal swab samples (data not shown) additional higher mass peaks greater than 80,000m/z could also be seen.

**Figure 4.**
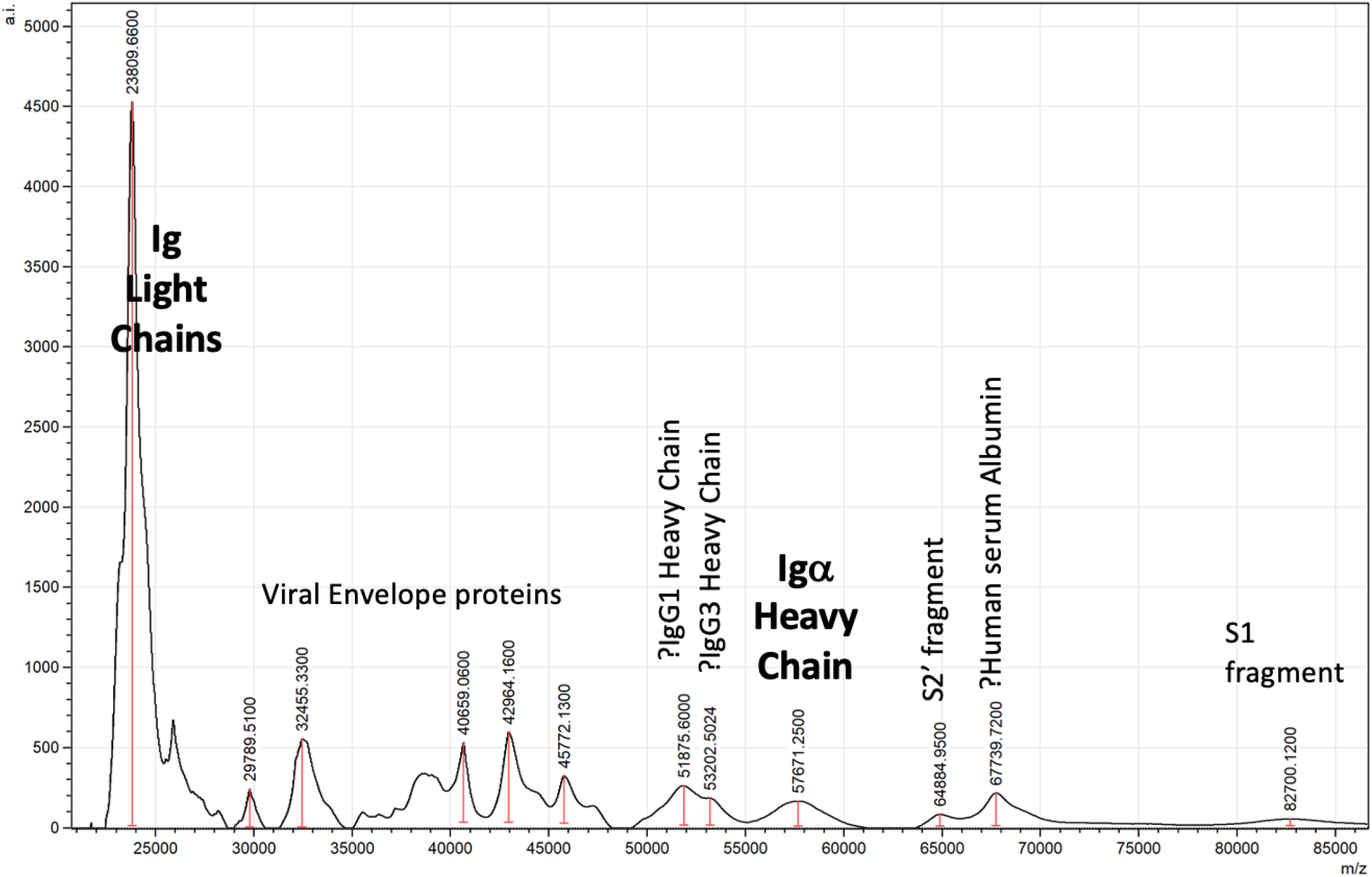
Example spectra of swab sample from SARS-CoV-2 RT-qPCR Brazilian patient with peak identification annotation after allowing for a 3-4% drift in higher mass calibration. S1 - Spike protein fragment 1; S2 - Spike protein fragment 2.

Given the drift in mass changes in mass and new peaks being found ML was used to discover any spectral pattern correlating with RT-qPCR positive swab samples. The ML system incorporated all 23 spectral features (peak presence and intensity) and the agreement with RT-qPCR was 78% for both random forest and extra tree algorithms and a strong correlation exists between both ML methods (Figure 5).

**Figure 5.**
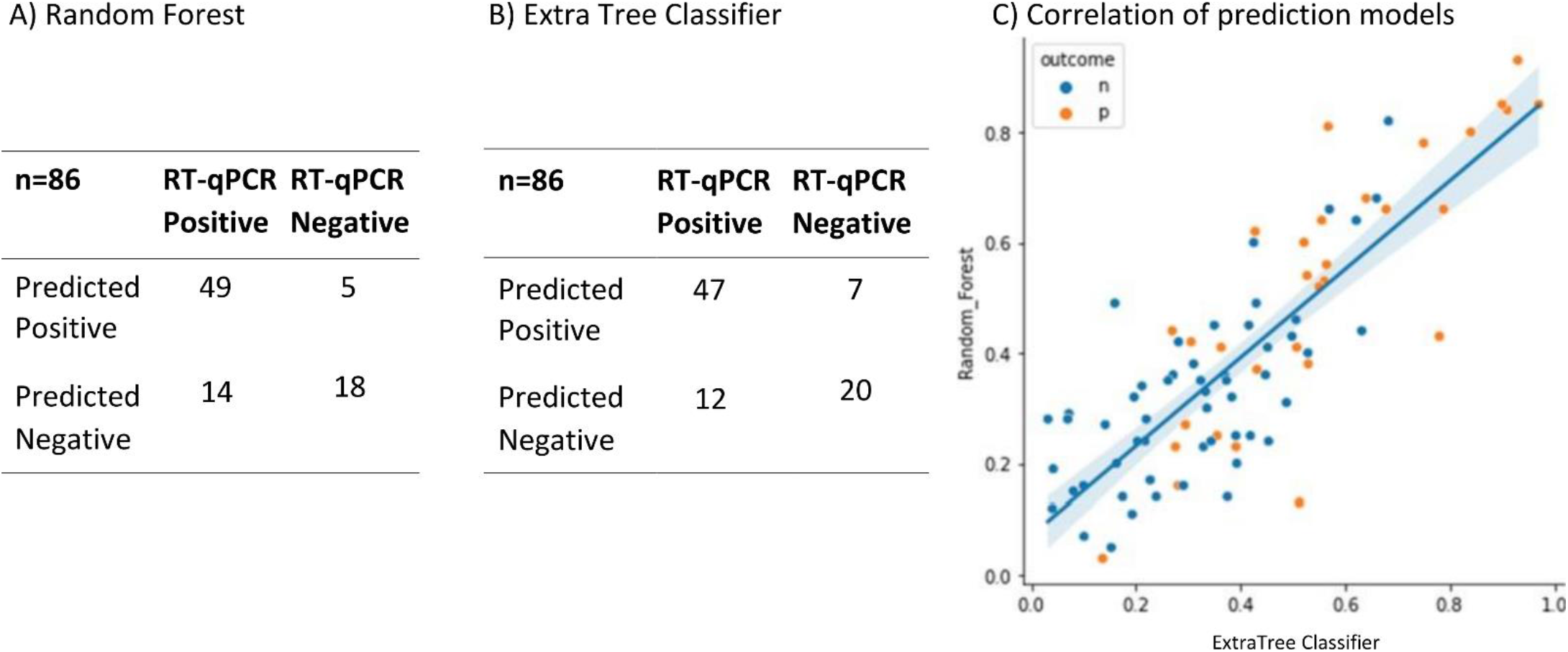
Performance characteristics of ML algorithms. Of the 215 sample spectra, 129 (60%) were randomly selected to form the training set. It was found that Random Forest (A) and Extra Tree Classifier (B) ML algorithms performed the best. A testing set of 86 samples (which had been independent from the training set) were run on the two algorithms and the results shown in the two confusion matrices (Panel A and B respectively). Agreement with RT-qPCR classification was 78%, p<0.00001 for both models. There was a strong correlation between both models on sample scoring for these validation test samples (panel C, line represents linear correlation r^2^ =0.681 and shading the 95% confidence limits. Blue dots (n) = negative, orange dots (p) = positive.

## 4. Discussion

The policy of RT-qPCR for SARS-CoV-2 testing varies among countries depending on the nature of the healthcare system. Within the UK and USA, community and asymptomatic surveillance testing programmes encouraged testing individuals who did not present with symptoms. Consequently, throughout the pandemic, 30-40% of positive RT-qPCR results for SARS-CoV-2 were reported in individuals not report having any symptoms [12]. This supported the arguments that RT-qPCR testing could detect pre-symptomatic and asymptomatic infections and therefore help control the spread of the disease (17). However, the definition of asymptomatic being RT-qPCR positive with no other clinical confirmation of infection, or subsequent development of symptoms, does become a self-serving metric; and does not account for any false positive finding in RT-qPCR testing (17). Among the Brazilian clinical samples analysed, 92.3% of the patients reported respiratory symptoms and RT-qPCR screening used only specific nucleocapsid (N) primers CDC: N1 e N2. Although in assessment studies no false positives were found when using these primers against *in vitro* cultures of other respiratory viruses; only 47% of nasopharyngeal samples from confirmed COVID-19 patients scored positive at CT<35. Whilst in leverage/sputum samples from the same patients 72% were positive [13,14].

The clinical laboratory protocol in Brazil was to re-test all samples with a Ct between 39 and 35 using CDC N1 and N2 primers, with the Seegene: E, RdRP, S e N primer reagent kit [13]. This simultaneously detects 3 target genes specific for SARS-CoV-2 in a single tube: RNA-dependent RNA polymerase, RdRP, N specific for SARS-CoV-2 and Envelope (E) for all sarbecovirus (including SARS-CoV-2): rarely is amplification seen in all target genes but 65% of COVID-19 patients nasopharyngeal samples showed amplification of at least two of the genes in evaluation trials [14]. Thus, two of three SARS-CoV-2 gene pairs being amplified (Ct <35) were regarded as being positive [14]. Therefore, re-evaluation of indeterminate results (i.e. Ct of 35-40 on the CDC: N1 e N2 RT-qPCR) with Seegene: E, RdRP, S e N was designed to improve detection rates without compromising specificity. Nevertheless, for the samples in the study this approach was not required, since all positive samples presented a Ct < 35 and negative samples presented a Ct > 40.

However, the self-serving definition of asymptomatic being RT-qPCR positive without ever developing symptoms, requires a complementary but orthogonal technology to RT-qPCR testing for SARS-CoV-2 infection detection. Given the costs of RT-qPCR testing it needed to be much more affordable and preferably to be a front-line testing system. Lateral flow devices that immunologically detect viral antigens has been introduced in many countries. However, this lacked sensitivity and only confirmed infection in those with symptomatic disease with extremely poor correlation with RT-qPCR pre-symptomatic and asymptomatic detection [15-17]. Mass spectrometry was proposed as a more sensitive method of antigen detection and we had built a system based on MALDI-TOF MS analysis for viral proteins [8]. This needed to be validated by other countries and on clinical samples.

The issues with travel restrictions during the pandemic has meant that emerging new technology from one country could only be replicated in another from written and verbal communication, and not via demonstration and in-person pedagogy. This had been the case of previously reported validation of our MALDI-TOF mass spectral analysis technique for SARS-CoV-2 infection based on gargle-saliva samples by a USA CLIA laboratory. This had specifically looked at SARS-CoV-2 viral infection in those presenting without symptoms [18]. For the Brazilian validation reported here, this was further complicated by not having identical MALDI-TOF mass spectrometers and the samples changing from gargle-saliva to nasopharyngeal swabs.

Given the underlying principles and preparation techniques of analysis remained true this should be possible. The initial hurdle was establishing the correct instrument settings for a Bruker Biotyper which would normally be installed and calibrated for relatively small bacterial culture ribosomal proteins and not large viral glycoproteins. This was achieved in the UK with direct comparison to the Shimadzu MALDI 8020 and a Bruker Biotyper loaned specifically for this purpose. The UK results showed extremely comparable data but greater sensitivity with respect to large viral glycoprotein detection by the Bruker Biotyper (See figures 1,2 & 3). However, initial calibration settings of the Bruker Biotyper instrument did later manifest as an added complication. Basically, the process and regression equation fit used in the Biotyper MALDI-TOF MS in Brazil (for bacterial identification and set by the manufacturers) may have provided reproducible and very precise calibration of the much smaller bacterial ribosomal proteins; but caused a drift of molecular mass overestimation at the much higher masses of viral glycoproteins being measured. In the UK all settings had been opened for change by the research team. However, in Brazil, as their instrument was routinely used for bacterial identification, this was not altered for the viral studies reported here. This could have been corrected in silico but was not, in case other bias and artefacts were introduced.

The processing of swabs rather than gargle-saliva samples presented only small changes in the overall preparation and was a matter of volume adjustments. However, several profile changes were evident. A Peak mass consistent with human serum albumin became a more prominent and constituent peak, as did two peaks consistent with the masses seen in other studies for Igγ1 and Igγ3 heavy chains [9]. This may reflect a more exudate induced difference between physical mucosal tissue abrasion of swabbing and a predominantly saliva and gentle mucous wash of a gargle.

In a smaller subset of the Brazilian swab samples broad peaks at around 90K and 100K were also detected and were similar in mass to those found in Lenti-viral pseudotype expressing coronavirus spike genes mass spectra (Figure 1).

Given the increased sensitivity of the Bruker Biotyper and subtle change in peak composition (and mass positions) a ML approach to differential analysis was adopted. For both of the ML algorithm systems adopted there was a 78% concordance with RT-qPCR scoring of infection.

Given that RT-qPCR is not 100% definitive of SARS-CoV-2 infection, or non-infection, this level of agreement strongly suggests that MALDI-TOF MS is indeed an orthogonal but complimentary screening test for SARS-CoV-2 infection. This is particularly valuable in pre-symptomatic and asymptomatic screening; for which lateral flow testing is not suitable, and has historically only identified no more than 4% of pre-symptomatic/ asymptomatic samples identified as positive by RT-qPCR [19]. Here 56-62% of RT-qPCR positives were also scored positive by MALDI-TOF Mass spectral profiling. Negative RT-qPCR agreement was 87-91%.

In conclusion MALDI-TOF mass spectral analysis is a complementary and orthogonal clinical test to RT-qPCR. Where agreement on SARS-CoV-2 infection is found on both tests, in patients presenting without symptoms, the diagnosis of infection as asymptomatic can be confidently made. In addition, where the test results diverge further analysis of clinical relevance is required as the potential to detect emergence of variants is a strong possibility.

## 5. Conclusions

MALDI-TOF mass spectral analysis is a complementary and orthogonal clinical test to RT-qPCR. Where agreement on SARS-CoV-2 infection is found on both tests, in patients presenting without symptoms, the diagnosis of infection as asymptomatic can be confidently made. Where the test results diverge further analysis of clinical relevance is required.

## Data Availability

All data produced in the present study are available upon reasonable request to the authors

## Supplementary Materials

Not applicable.

## Author Contributions

For research articles with several authors, a short paragraph specifying their individual contributions must be provided. The following statements should be used “Conceptualization, Lovison, O. A.; Grigaitė, R.; Volpato, F. C. Z.; Iles, J. K.; Lacey, J.; Barreto, F.; Balzan, L. L. R.; Cantarelli, V. V.; Barth, A. L.; Martins, A. F.; Iles, R. K.; methodology, Lovison, O. A.; Grigaitė, R.; Volpato, F. C. Z.; Iles, J. K.; Lacey, J.; Iles, R. K; software, Grigaitė, R; Pandiri, S. R.; validation, Lovison, O. A.; Grigaitė, R.; Volpato, F. C. Z.; Pandiri, S. R.; Martins, A. F.; Iles, R. K.; formal analysis, Lovison, O. A.; Grigaitė, R.; Martins, A. F.; Iles, R. K.; investigation, Lovison, O. A.; Grigaitė, R.; Martins, A. F.; Iles, R. K.; resources, Barreto, F.; Balzan, L. L. R.; Cantarelli, V. V.; Barth, A. L.; Martins, A. F.; Iles, R. K.; data curation, Lovison, O. A.; Grigaitė, R.; Pandiri, S. R.; Iles, R. K.; writing—original draft preparation, Lovison, O. A.; Grigaitė, R.; Iles, R. K; writing—review and editing, Martins, A. F.; Iles, R. K.; visualisation, Martins, A. F.; Iles, R. K.;; supervision, Martins, A. F.; Iles, R. K.; project administration, Martins, A. F.; Iles, R. K.; funding acquisition, Barreto, F.; Barth, A. L.; Martins, A. F.; Iles, R. K.; All authors have read and agreed to the published version of the manuscript.” Please turn to the [CRediT taxonomy](https://img.mdpi.org/data/contributor-role-instruction.pdf) for the term explanation. Authorship must be limited to those who have contributed substantially to the work reported.

## Institutional Review Board Statement

The study was conducted in accordance with the Declaration of Helsinki, and approved by the Ethics Committee of HOSPITAL DE CLÍNICAS DE PORTO ALEGRE (protocol code 4.355.906 and 22 Oct. 2020).

## Informed Consent Statement

Informed consent was obtained from all subjects involved in the study.

## Data Availability Statement

The data presented in this study are available on request from the corresponding author. The data are not publicly available due to being used for further investigation and developments.

## Acknowledgments

Not applicable.

